# Swab-based molecular testing near the point-of-care for childhood tuberculosis: a prospective diagnostic accuracy study

**DOI:** 10.64898/2026.11.16.26368102

**Authors:** Devan Jaganath, Aggrey Ainebyona, Juaneta Luiz, Hayley Poore, Denise Banze, Peter Wambi, Robert Castro, Seda Yerlikaya, Payam Nahid, Claudia M. Denkinger, Adithya Cattamanchi, Celso Khosa, Heather J. Zar, Nilesh Bhatt, Eric Wobudeya, the R2D2 TB Network and ADAPT for Kids Studies

## Abstract

**Background:** Swab-based molecular testing for tuberculosis (TB) could provide a near point-of-care (nPOC) diagnostic for children. We evaluated the accuracy of two swab-based nPOC platforms to detect pediatric pulmonary TB.

**Methods:** We prospectively enrolled children under 15 years with presumptive TB in Uganda and South Africa. They completed TB evaluation including sputum molecular testing and culture. Two tongue swabs were collected and tested with MiniDock MTB (Pluslife Biotech, China) and Truenat MTB Ultima (Molbio Diagnostics, India), and one swab was dipped in sputum for testing with MiniDock MTB. We calculated the sensitivity and specificity of each index test against a culture-based standard and compared them with sputum molecular testing and smear microscopy.

**Findings:** We included 335 children, among whom median age was 3 years (IQR 1-6), 53% were female, 11% were living with HIV, and 16% had microbiologically confirmed TB. Tongue swabs were collected in all children, with invalid test rates of less than 3%. Against a culture-based standard, the sensitivities of tongue swab MiniDock MTB and MTB Ultima were 50% (95% CI 33.4-66.6) and 50% (95% CI 28.2-71.8), respectively, and specificities were ≥98% for both. Sputum swab MiniDock MTB had similar sensitivity to sputum Xpert Ultra (85.7% vs. 75.0%, p=0.45), but substantially higher sensitivity than tongue swab MiniDock MTB (+23.1%, p=0.03) and sputum smear microscopy (+46.4%, p<0.001).

**Interpretation:** Sputum swab testing had similar accuracy to sputum Xpert Ultra in children. Tongue swab testing had moderate accuracy, and has value to identify microbiologically confirmed cases when sputum is not available.

**Funding:** National Institutes of Health, United States Department of State Bureau of Global Health Security and Diplomacy, and South African Medical Research Council

**RESEARCH IN CONTEXT:** *Evidence before this study:* We evaluated the literature from October 2021 to October 2026 for studies in any language on oral swab molecular testing for *M. tuberculosis* (MTB). We included diagnostic accuracy studies and excluded work on extrapulmonary TB, screening and early assay development, and searched PubMed and Google Scholar with the terms “swab” AND “tuberculosis” AND “diagnosis.” Manual quantitative polymerase chain reaction (qPCR) testing on tongue swabs has high accuracy to detect pulmonary TB in adults (80-93% sensitive and >98% specificity), with similar accuracy to sputum samples. Translation to semi-automated PCR assays such as Xpert Ultra have had high specificity but variable sensitivity (59-76%). Newer assays have been developed that are designed specifically for swab testing, and are small, portable, and can be used near the point-of-care (nPOC). Studies in adults have found that nPOC tongue swab testing is feasible, sensitive (75-86%), and specific (>96%). Testing can also be performed by dipping a swab in sputum, offering higher accuracy than tongue swabs and similar performance to sputum Xpert Ultra. However, the evidence in children has been limited to Xpert Ultra tongue swab testing, with low sensitivity (7-19%).

*Added value of this study:* To our knowledge, this is the first assessment of two swab-based nPOC assays for childhood TB: MiniDock MTB and MTB Ultima. In two high TB-burden settings of South Africa and Uganda, we prospectively enrolled children with presumptive TB and determined the accuracy of tongue and sputum swab testing. We found that tongue swab testing detected about half of children with culture-confirmed TB; sputum swab testing had substantially higher sensitivity than tongue swabs, and similar performance to sputum Xpert Ultra, meeting the World Health Organization (WHO) target accuracy for an nPOC sputum molecular TB test.

*Implications of all the available evidence:* The recent WHO guidelines endorsed sputum swab testing in adults and adolescents, and tongue swabs if sputum is not available. Similarly, we found that a single platform can enable nPOC molecular testing for TB among children, with sputum swab accuracy comparable to Xpert Ultra when sputum is available, and tongue swabs providing a simple, non-invasive approach to increase microbiological confirmation and potentially reduce delays in treatment when sputum is not available.

## INTRODUCTION

Children have limited access to tuberculosis (TB) diagnostics worldwide. Standard sputum-based molecular testing for *Mycobacterium tuberculosis* (MTB) is challenging, as young children are frequently unable to expectorate and require sputum induction or gastric aspiration, which are more invasive and may not be available at lower-level facilities.^1^ Referral to higher-level facilities creates additional access barriers such as transportation needs and greater costs.^2^ Consequently, only half of the estimated 1.2 million children with TB disease are reported to public health programs,^3^ suggesting massive underdiagnosis. There is an urgent need for non-sputum tests that can be implemented at or near the point-of-care (nPOC).^4^

Tongue swab-based assays have greatly advanced, enabling simpler, faster, low-cost molecular detection, with several novel nPOC molecular devices developed during the COVID-19 pandemic.^5^ nPOC devices are now recommended by the World Health Organization (WHO) for use in adults and adolescents, including patients with smear-negative TB.^6^ They are also more sensitive than smear microscopy, preferred over sputum testing, and are cost-effective.^7–10^ If sputum can be obtained, the same platform can be used by dipping the swab in sputum, offering diagnostic accuracies similar to those of sputum Xpert Ultra.^7,10^

Sputum swab testing was extrapolated to children with limited data, but tongue swab testing was not endorsed for children. Prior efforts in children using tongue swabs with the GeneXpert (Cepheid, Sunnyvale USA) platform have been low yield;^11,12^ however, the Xpert MTB/RIF and Xpert MTB/RIF Ultra (Xpert Ultra; Cepheid) cartridges were designed and optimized for sputum. Thus, we prospectively assessed the accuracy of two nPOC platforms, MTB Nucleic Acid Test Card (MiniDock MTB, Pluslife Biotech, Guangzhou, China) and Truenat MTB Ultima (MTB Ultima; Molbio Diagnostics, Goa, India), on tongue swabs and on sputum swabs to detect pulmonary TB in children in Uganda and South Africa.

## METHODS

### Study Design and Participants

We conducted a prospective diagnostic accuracy study as part of the R2D2 Kids and ADAPT for Kids studies (ClinicalTrials.gov registration #NCT05989802). We recruited children from October 3, 2024 to December 11, 2025 who had symptoms or risk factors for pulmonary TB in Kampala, Uganda from Mulago National Referral Hospital and surrounding Kampala Capital City Authority and Infectious Diseases Institute clinics and hospitals, and in Gqeberha, South Africa, at Dora Nginza Hospital with referrals from surrounding outpatient clinics from the Nelson Mandela Bay district. We included children if they had two or more of the following: 1) Unexplained cough for any duration; 2) Known TB contact or tuberculin skin test or interferon gamma release assay positive; or 3) Abnormal chest X-ray (any abnormality). If they had only one of these, they could be included if they had also experienced a) Unexplained weight loss, unexplained failure to thrive, or severe acute malnutrition; b) Unexplained fever for 2 weeks or longer; or c) Unexplained lethargy or reduced playfulness for 2 weeks or longer. We excluded children if they completed treatment for TB disease or infection in the last year, were on anti-TB medication for more than 3 days, or were unable to return for follow-up visits. All adult caregivers provided written informed consent, and capable children aged 8 years and older in Uganda or 7 years and older in South Africa provided written informed assent. The study was approved by the Human Research Ethics committee of the Faculty of Health Sciences, University of Cape Town, the Mulago Hospital Ethical Research Committee, the Uganda National Council on Science and Technology, and the University of California, San Francisco Institutional Review Board.

### TB Evaluation Procedures

We performed standard TB diagnostic procedures as defined by consensus guidelines for TB diagnostic studies in children.^13–15^ All children completed a standardized history and physical examination, and routine TB evaluation included HIV testing, two-view chest X-ray (CXR), and collection of two respiratory specimens at least four hours apart (ie, induced sputum, gastric aspirates, or nasopharyngeal aspirates). Stool was collected if a respiratory specimen was not possible or unsuccessfully collected. HIV testing and interpretation followed each site’s national guidelines. CXRs were independently read by two physicians with expertise in the evaluation of childhood TB, and classified as Abnormal - Consistent with TB, Abnormal – Equivocal, or Normal. A third physician read the CXR as a tiebreaker, if needed.

A molecular WHO-recommended rapid diagnostic (mWRD), acid-fast bacilli (AFB) smear microscopy, and culture testing were performed on the respiratory samples. In Uganda, the standard mWRD is Xpert Ultra; in South Africa, both Xpert Ultra and Cobas MTB-RIF/INH (Roche, Basel, Switzerland) were used. Manufacturer instructions were followed for both, with a result output of positive (MTB detected), negative (MTB not detected), or invalid. If both samples were invalid, Xpert Ultra testing was repeated. If stool was collected, Xpert Ultra testing was performed using the simple one-step (SOS) processing method.^16^ Positive Xpert Ultra results also provide a semi-quantitative output, and trace was defined as positive in children as per WHO guidance.^17^ Dedicated mycobacteriology labs performed fluorescent smear microscopy with Auramine-O staining, and mycobacterial culture using liquid (Mycobacterial Growth Indicator Tube (MGIT), 37°C for 6 weeks), or solid media (Löwenstein–Jensen (LJ), 37°C for 8 weeks). MTB was confirmed using an MPT64 TB Ag kit (SD Biosensor, Suwon-si, Republic of Korea).

Initiation of TB treatment was determined by local clinicians according to the standard of care, without access to swab test results. All children, regardless of TB status, had follow up evaluations at 2-4 weeks and 2-3 months to assess clinical status and any response to treatment. If anti-TB medication was initiated at a follow up visit, they returned for an additional assessment 2-3 months later.

### Swab Collection

Two separate tongue swabs were obtained for MiniDock MTB and MTB Ultima testing, alternating the order of testing of each assay weekly. A Copan FLOQswab was applied to the dorsal tongue in a back-front and left-right motion for 30 seconds with a timer, focusing on the back of the tongue and rotating the swab throughout. For MiniDock MTB, the swab head was placed into a vial pre-filled with a buffer solution, swirling 10 times and then pinching the swab by squeezing the outside of the tube as the swab was removed. The swab was removed and the vial sealed. For MTB Ultima, the swab head was also placed in a pre-filled vial, but the head was broken off and retained in the vial after swirling.

Sputum swab preparation for MiniDock MTB testing was performed in Uganda, prior to mWRD testing. A Copan FLOQswab was immersed into the sputum sample and swirled ten times for 15 seconds using a timer. The sputum sample included expectorated or induced sputum, or gastric or nasopharyngeal aspirates. The swab was placed in the pre-filled vial with the same procedure as the MiniDock MTB tongue swab.

### Swab testing

Fresh samples were kept at ambient temperature and tested within 24 hours according to manufacturer instructions. In brief, for MiniDock MTB, the sample was heated to 75°C and lysed using the Pluslife Thermolyse device after five minutes. The lysate was squeezed into a Nucleic Acid Test Card (Reaction Card) until the liquid was between two indicated injection lines. The sealed reaction card stood for 15 seconds, and then was shaken up and down ten times for five seconds. The reaction card was then placed in an Integrated Nucleic Acid Testing Device to begin testing. MiniDock uses RNase Hybridization Assisted Amplification (RHAM) technology for isothermal amplification. After running for approximately 25 minutes, an indicator light showed a positive, negative, or invalid result. If invalid, the test was repeated with the remaining lysate. This procedure was performed for both tongue and sputum swabs. Prior to the study, every 100 cards, and at completion, a positive and negative control was tested to ensure the device was functioning appropriately.

For MTB Ultima, the sample was heated and lysed using the Molbio Truelyse device for 90 seconds. Six microliters of the lysate were then added to a pre-filled PCR reagent tube, incubated for 30-60 seconds, and then added to the MTB Ultima chip. The chip was tested in a Molbio Truelab device for 40 minutes, and the result was then displayed on the screen as positive, negative, or invalid. If invalid, the test was repeated with the remaining lysate.

The laboratory staff was blinded to the TB status of the participant when performing swab testing. To reduce cross-contamination with sputum testing, the swab processing was performed separately in a biosafety cabinet. An air swab was obtained from specimen collection areas (swirling a swab in the air for 30 seconds) weekly as well as for any participant result that was swab positive, but sputum mWRD negative. The air swab was then tested using Xpert Ultra per a consensus protocol.^18^

### Reference Standard

The primary reference standard was based on mycobacterial culture, in order to align with the WHO target specification standard (TSS)-17 for *in vitro* diagnostic medical devices for qualitative detection of MTB DNA.^19^

Participants were also classified as Confirmed, Unconfirmed, or Unlikely TB per NIH consensus definitions.^15^ Confirmed TB was defined as having molecular or culture positivity for MTB on at least one respiratory sample. Unconfirmed TB was classified if microbiological testing was negative, but there was other evidence of TB disease including at least two of: symptoms and signs suggestive of TB, a close TB contact or immunological evidence of TB infection, CXR consistent with TB, and/or the child was started on anti-TB treatment with improvement or resolution in symptoms. Unlikely TB cases had symptoms of lower respiratory tract illness, but did not have microbiological evidence of MTB, did not meet Unconfirmed TB criteria, and improved without anti-TB treatment. Secondary reference standards included: 1) A microbiological reference standard (MRS) defined as Confirmed TB vs. Unconfirmed or Unlikely TB; and 2) A composite reference standard (CRS) defined as Confirmed or Unconfirmed TB vs. Unlikely TB.

### Statistical Analysis for Diagnostic Accuracy

Sample size was determined by a planned enrollment period of approximately one year. Sensitivity, specificity, positive predictive values (PPVs), and negative predictive values (NPVs) were calculated for tongue swab and sputum swab testing with 95% confidence intervals (CIs) against the culture-based reference standard, MRS, and CRS. Children with missing or persistently invalid index or reference test results were excluded from the analysis. The primary benchmark accuracy was the WHO target product profile (TPP) for an nPOC molecular TB test for sputum (≥85% sensitivity and ≥98% specificity) or non-sputum (≥75% sensitivity and ≥98% specificity) samples.^4^ At the same time, it is important to recognize that there are WHO-approved sample types for molecular TB testing in children that do not meet this TPP accuracy, including nasopharyngeal aspirate (NPA). Thus, we also evaluated a contextual benchmark based on the accuracy of NPA for childhood TB, at 46% sensitivity and 98% specificity against culture.^20^ We calculated the proportion of invalid results for each test, with a goal invalid proportion of <5% per the WHO TPP. Accuracy was also determined by key subgroups, including age category, respiratory sample type, HIV status, and underweight status (WHO weight-for-age Z score <-2 if age under 10 years or body mass index (BMI)-for-age Z-score <-2 if age 10-14 years).^21^

Across each reference standard, we compared tongue swab MiniDock MTB, tongue swab MTB Ultima, and sputum swab MiniDock MTB to sputum mWRD and sputum smear microscopy. For sputum swab versus sputum mWRD and smear, we compared the results from the same sample to reduce bias with multiple testing. For tongue swab versus sputum mWRD and smear, we used the sputum collected closest to the time of tongue swab collection. We calculated the absolute percentage difference using McNemar’s test, with significance defined as p-value < 0.05. Stata version 18.5 (Statacorp, College Station, TX, USA) software was used. Results were reported according to the Standards for Reporting of Diagnostic Accuracy Studies (STARD) criteria.^22^

### Role of the funding source

The funders did not have a role in the in study design, collection, analysis, and interpretation of data, in the writing of the report, or in the decision to submit the paper for publication.

## RESULTS

### Cohort Characteristics

The flow of participant enrollment is shown in Figure 1. Of 335 participants enrolled, all provided a respiratory sample (>80% induced sputum) and a tongue swab. The median age was 3 years (IQR 1-6), 53.4% were female, 10.5% were living with HIV, and 24.8% were underweight (Table 1).

**Figure 1.**
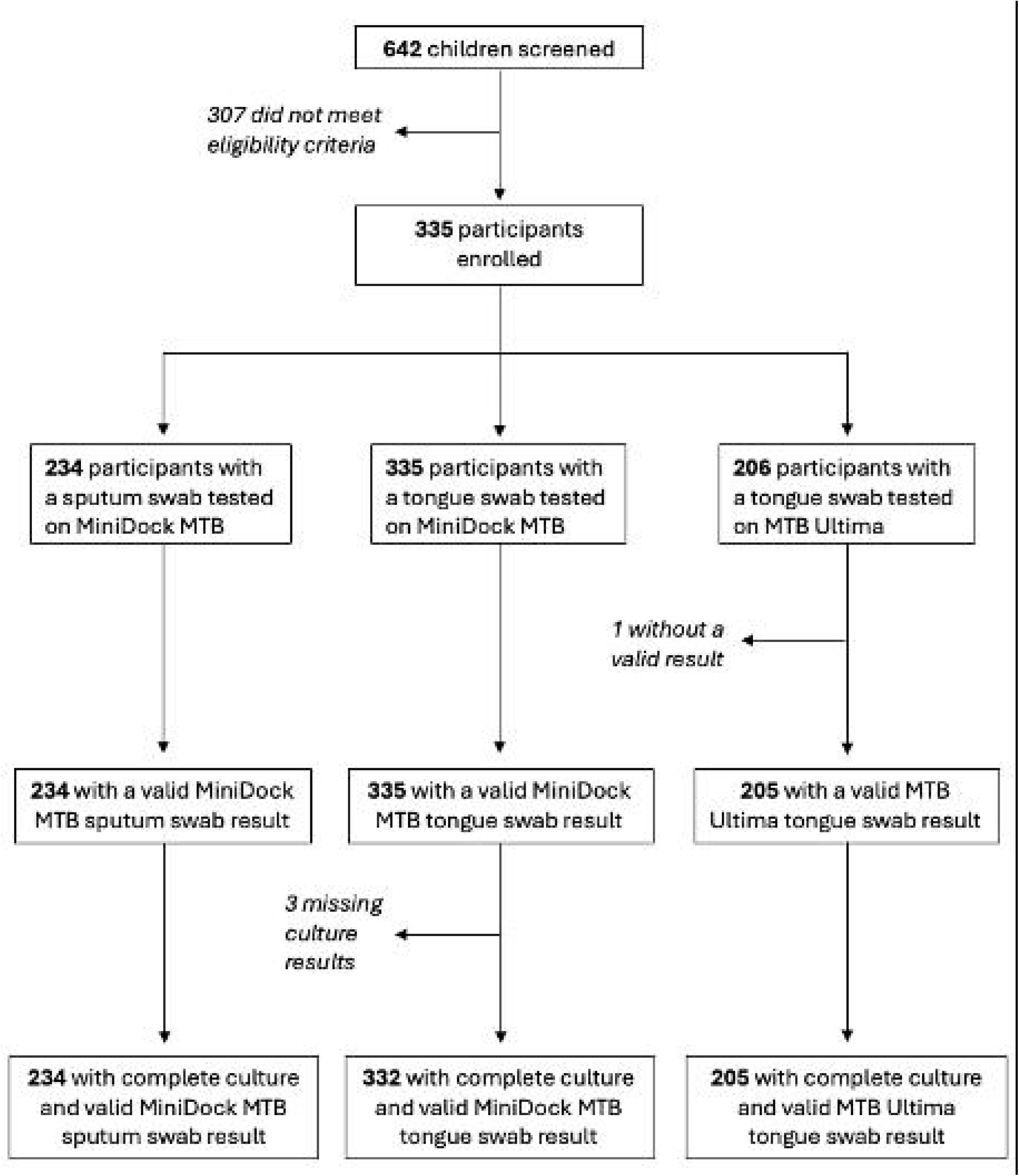
Participant flowchart.

**Table 1.**
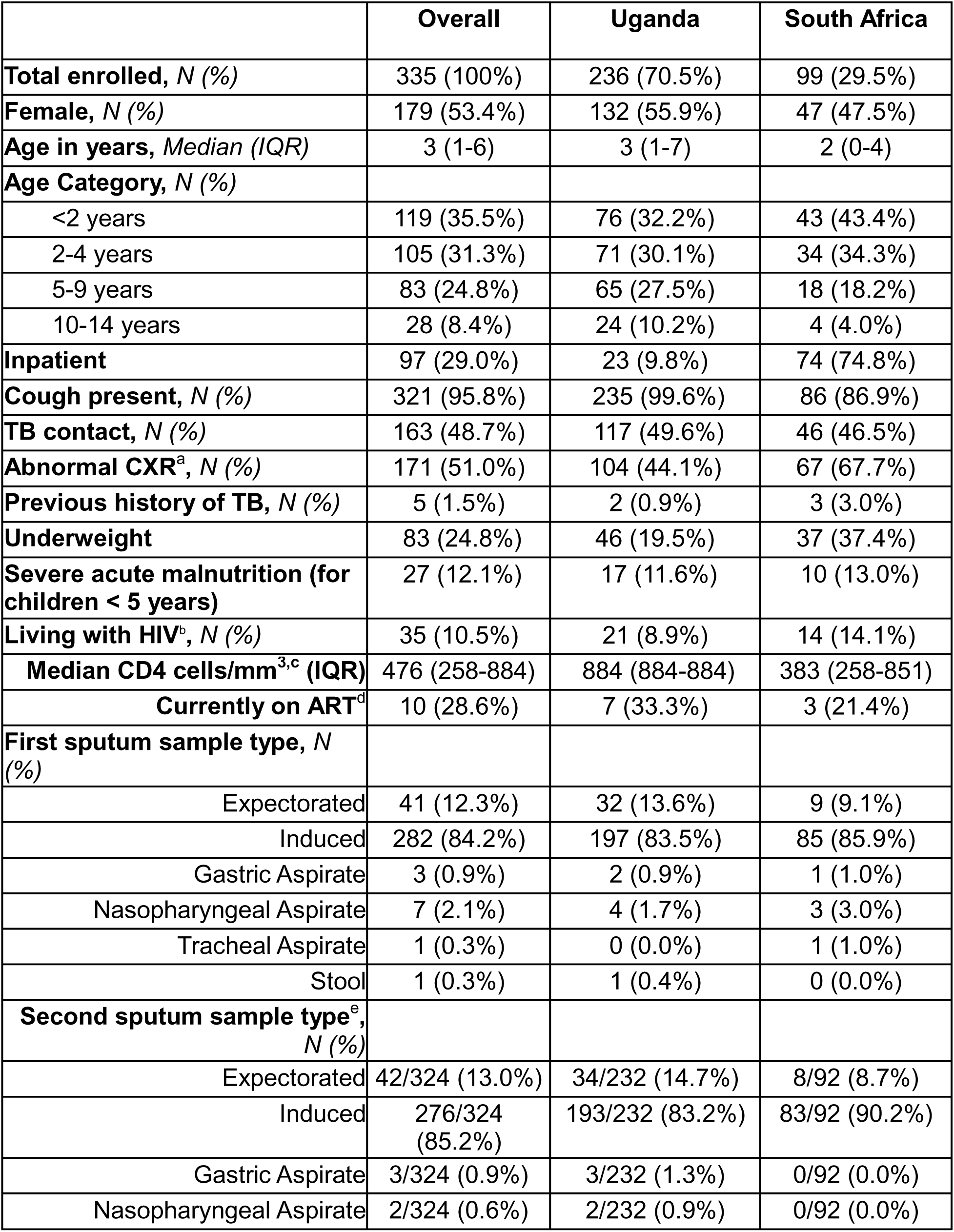
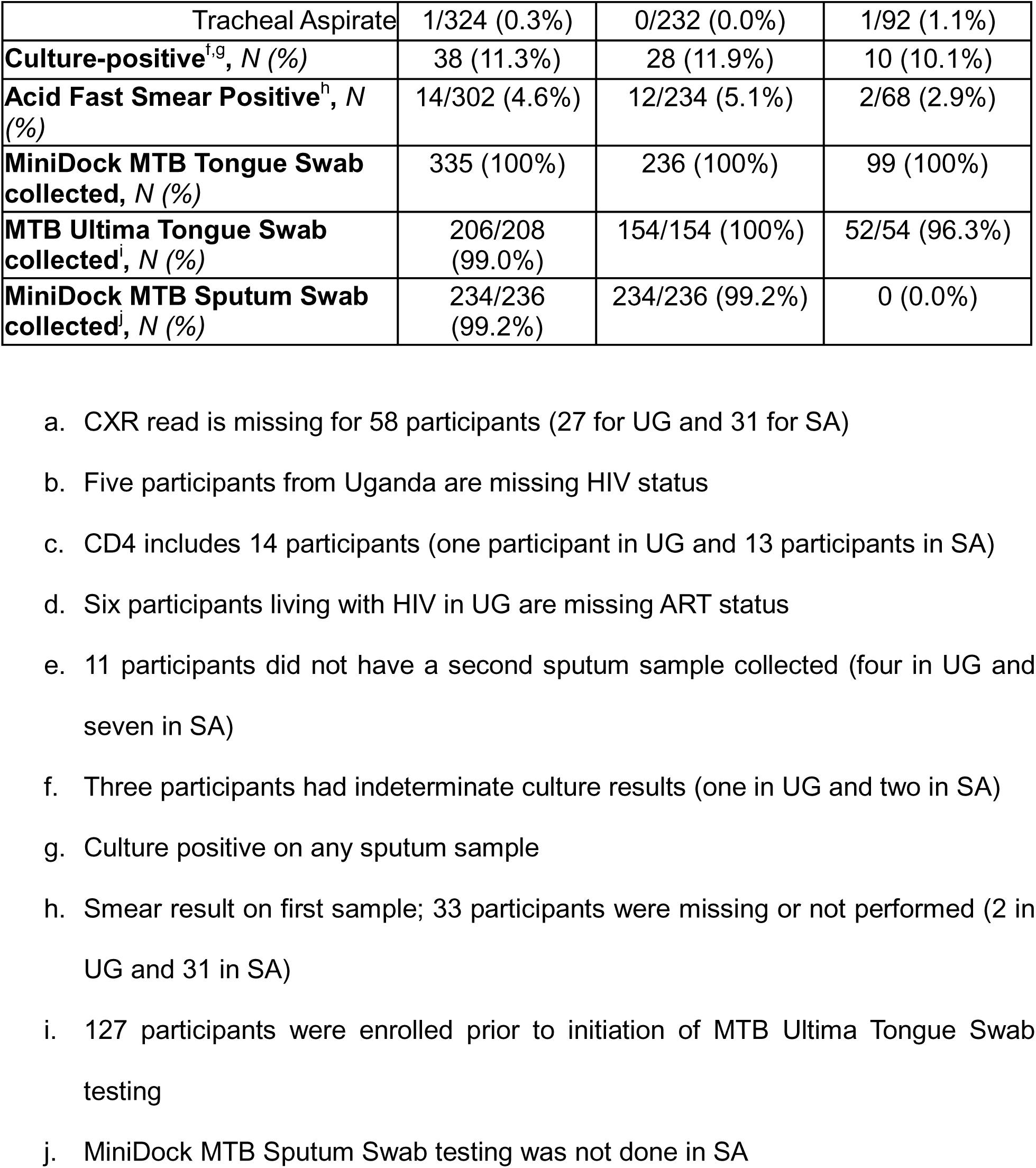
Participant characteristics, overall and by country. a. CXR read is missing for 58 participants (27 for UG and 31 for SA) b. Five participants from Uganda are missing HIV status c. CD4 includes 14 participants (one participant in UG and 13 participants in SA) d. Six participants living with HIV in UG are missing ART status e. 11 participants did not have a second sputum sample collected (four in UG and seven in SA) f. Three participants had indeterminate culture results (one in UG and two in SA) g. Culture positive on any sputum sample h. Smear result on first sample; 33 participants were missing or not performed (2 in UG and 31 in SA) i. 127 participants were enrolled prior to initiation of MTB Ultima Tongue Swab testing j. MiniDock MTB Sputum Swab testing was not done in SA

Overall, 38 (11.3%) were culture positive, representing 38/195 (19.5%) of children diagnosed with TB. By NIH classification, 54 (16.1%) had Confirmed TB, 141 (42.1%) had Unconfirmed TB, and 124 (37.0%) had Unlikely TB. By country, children from South Africa were younger (43% under 2 years) and more likely to be living with HIV (Table 1).

### Accuracy of tongue and sputum swab MTB testing

The accuracy of swab-based TB testing against a culture-based reference standard is shown in Table 2. The number and proportion of invalid results for the first tongue swab MiniDock MTB, MTB Ultima, and sputum swab MiniDock MTB were 4/335 (1.2%), 6/206 (2.9%), and 3/234 (1.3%), respectively. When the invalid samples were retested, all MiniDock MTB results were then valid and MTB Ultima had one invalid result. Of 332 tongue swabs with valid MiniDock MTB and culture results, the sensitivity was 50.0% (95% CI 33.4-66.6) and specificity was 98.3% (95% CI 96.1-99.4). Of 205 tongue swabs tested with MTB Ultima, the sensitivity was 50.0% (95% CI 28.2-71.8) and specificity was 97.8% (95% CI 94.5-99.4). Sputum swab MiniDock MTB testing was 85.7% sensitive (95% CI 67.3-96.0) and 98.1% specific (95% CI 95.1-99.5). The PPVs ranged from 73.3-79.2% for tongue swabs and 85.7% for MiniDock MTB, and NPVs ranged from 93.8-94.2% for tongue swabs and 98.1% for sputum swab MiniDock MTB (Table 3). In subgroup analysis, sensitivity was lower in children <5 years, and higher in those with HIV (Table 2).

**Table 2.** Diagnostic accuracy of tongue swab and sputum swab TB testing according to a sputum culture-based reference standard.

|  | MiniDock MTB Tongue Swab<br>N=332 | MTB Ultima Tongue Swab<br>N=205 | MiniDock MTB Sputum Swab<br>N=234 |
|---|---|---|---|
| Sensitivity<br>n/N, % (95% CI) | 19/38,<br>50.0% (33.4-66.6) | 11/22,<br>50.0% (28.2-71.8) | 24/28,<br>85.7% (67.3-96.0) |
| <5 years | 6/19,<br>31.6% (12.6-56.6) | 3/11,<br>27.3% (6.0-61.0) | 10/12,<br>83.3% (51.6-97.9) |
| Living with HIV | 2/3,<br>66.7% (9.4-99.2) | 1/1,<br>100% (2.5-100) | 2/2,<br>100% (15.8-100) |
| Underweight | 7/13,<br>53.8% (25.1-80.8) | 2/5,<br>40.0% (5.3-85.3) | 9/9,<br>100% (66.4-100) |
| Specificity<br>n/N, % (95% CI) | 289/294,<br>98.3% (96.1-99.4) | 179/183,<br>97.8% (94.5-99.4) | 202/206,<br>98.1% (95.1-99.5) |
| <5 years | 200/203,<br>98.5% (95.7-99.7) | 123/123,<br>100% (97.0-100) | 132/134,<br>98.5% (94.7-99.8) |
| Living with HIV | 32/32,<br>100% (89.1-100) | 20/21,<br>95.2% (76.2 -99.9) | 18/18,<br>100% (81.5-100) |
| Underweight | 68/69,<br>98.6% (92.2-100) | 42/44,<br>95.5% (84.5-99.4) | 37/37,<br>100% (90.5-100) |

**Table 3.** Diagnostic Accuracy of MiniDock MTB and MTB Ultima testing, by reference standard. CRS: Composite Reference Standard; MRS: Microbiological Reference Standard a. Three participants with a valid MiniDock MTB tongue swab result had an indeterminate culture result b. 16 participants did not have a CRS classification

| **Reference standard** | **MiniDock MTB Tongue Swab<br>N=335** | **MTB Ultima Tongue Swab<br>N=205** | **MiniDock MTB Sputum Swab<br>N=234** |
|---|---|---|---|
| **Sensitivity**<br>n/N, % (95% CI) |  |  |  |
| Culture<sup>a</sup> | 19/38,<br>50.0% (33.4-66.6) | 11/22,<br>50.0% (28.2-71.8) | 24/28,<br>85.7% (67.3-96.0) |
| MRS | 21/54,<br>38.9% (25.9-53.1) | 13/32,<br>40.6% (23.7-59.4) | 27/42,<br>64.3% (48.0-78.4) |
| CRS<sup>b</sup> | 23/195,<br>11.8% (7.6-17.2) | 14/126,<br>11.1% (6.2-17.9) | 28/126,<br>22.2% (15.3-30.5) |
| **Specificity**<br>n/N, % (95% CI) |  |  |  |
| Culture<sup>a</sup> | 289/294,<br>98.3% (96.1-99.4) | 179/183,<br>97.8% (94.5-99.4) | 202/206,<br>98.1% (95.1-99.5) |
| MRS | 278/281,<br>98.9% (96.9-99.8) | 171/173,<br>98.8% (95.9-99.9) | 191/192,<br>99.5% (97.1-100) |
| CRS<sup>b</sup> | 123/124,<br>99.2% (95.6-100) | 72/73,<br>98.6% (92.6-100) | 98/98,<br>100% (96.3-100) |
| **Positive predictive value**<br>n/N, % (95% CI) |  |  |  |
| Culture<sup>a</sup> | 19/24,<br>79.2% (57.8-92.9) | 11/15,<br>73.3% (44.9-92.2) | 24/28,<br>85.7% (67.3-96.0) |
| MRS | 21/24,<br>87.5% (67.6-97.3) | 13/15,<br>86.7% (59.5-98.3) | 27/28,<br>96.4% (81.7-99.9) |
| CRS<sup>b</sup> | 23/24,<br>95.8% (78.9-99.9) | 14/15,<br>93.3% (68.1-99.8) | 28/28,<br>100% (87.7-100) |
| **Negative predictive value**<br>n/N, % (95% CI) |  |  |  |
| Culture<sup>a</sup> | 289/308,<br>93.8% (90.5-96.2) | 179/190,<br>94.2% (89.9-97.1) | 202/206,<br>98.1% (95.1-99.5) |
| MRS | 278/311,<br>89.4% (85.4-92.6) | 171/190,<br>90.0% (84.8-93.9) | 191/206,<br>92.7% (88.3-95.9) |
| CRS<sup>b</sup> | 123/295,<br>41.7% (36.0-47.6) | 72/184,<br>39.1% (32.0-46.6) | 98/196,<br>50.0% (42.8-57.2) |

b. 16 participants did not have a CRS classification

### Head-to-head comparison

The head-to-head accuracy comparisons of swab-based testing, sputum mWRD, and smear microscopy are shown in Figure 2 and Supplemental Table 1. Tongue swab MiniDock MTB and MTB Ultima sensitivity was lower than sputum mWRD, but tongue swab MiniDock MTB was more sensitive than sputum smear microscopy (61.5% versus 30.8%, respectively, absolute difference 30.8%, 95% CI 9.2-52.4, p < 0.01). On sputum, MiniDock MTB swab had higher sensitivity than both tongue swab tests, and similar sensitivity to sputum Xpert Ultra. Specificity was high (≥97%) and similar across platforms. There were no positive air swab results during the study period. Tongue swab MiniDock MTB results were 95.7% concordant with sputum MiniDock MTB (Supplemental Table 2). Sputum swab had eight positive results that were not detected by tongue swab, and two tongue swab MiniDock MTB tests were not detected by sputum swab. Thus, there was an added yield of 10/234 (4%) if both sputum swab and tongue swab were performed.

**Figure 2.**
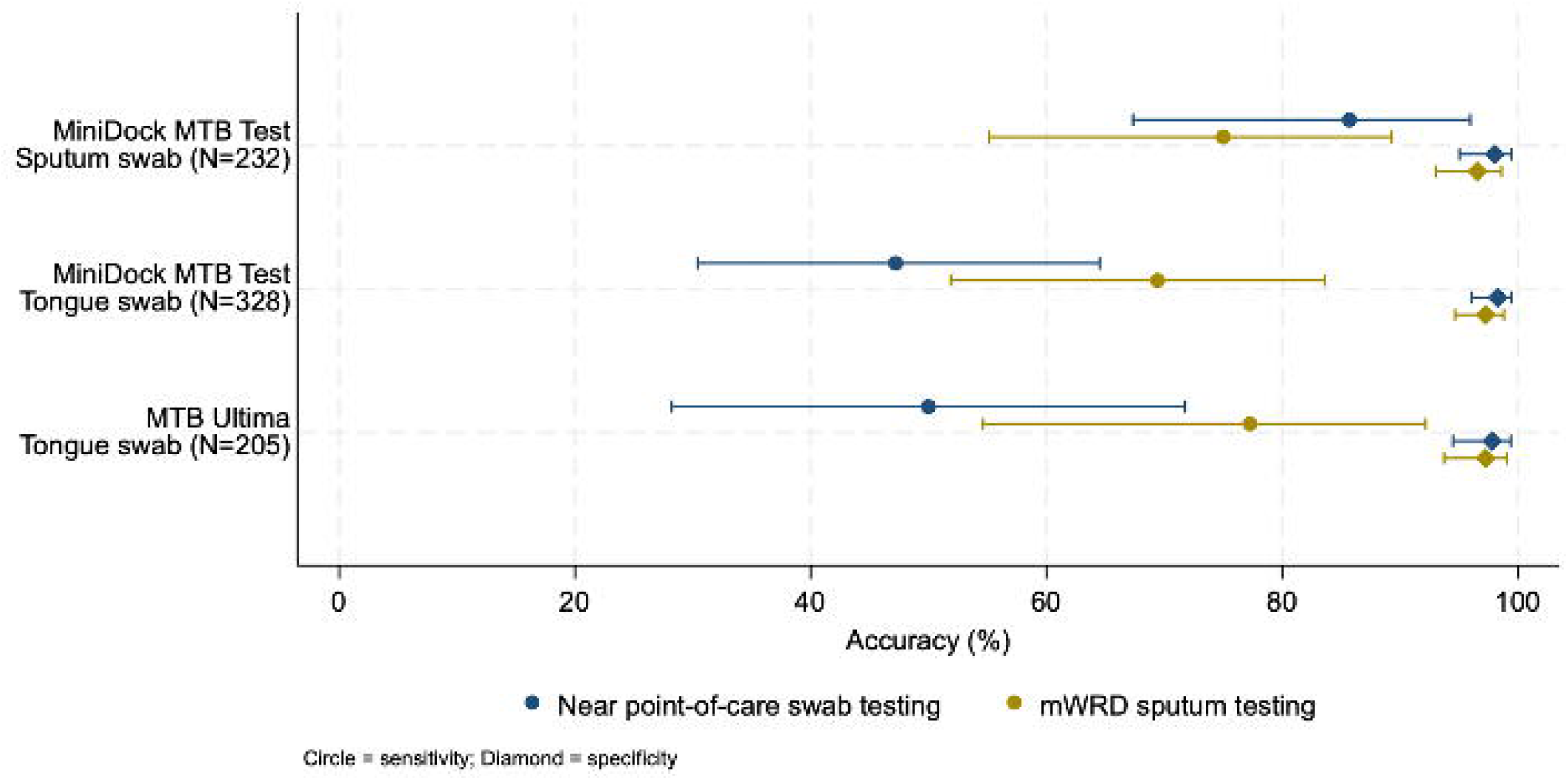
The head-to-head accuracy of near point-of-care swab testing compared to sputum mWRD testing. Sputum molecular results were compared from the same sputum specimen used for the sputum-swab index test, or from the sputum specimen with the closest collection date to the tongue-swab index test. mWRD: molecular WHO-recommended rapid diagnostic.

### NIH reference standards

Table 3 shows the diagnostic accuracy of tongue and sputum swab testing against the MRS and CRS. When sputum mWRD testing was included in the MRS definition of TB, the sensitivities of tongue swab testing reduced to 38.9% and 40.6% for MiniDock MTB and MTB Ultima, respectively. The sensitivities of tongue swab testing further reduced with the CRS to 11.8% and 11.1%. Sputum swab sensitivities also reduced with the MRS and the CRS as noted in Table 3. Specificity remained high at ≥98% for tongue swabs and sputum swab testing against all three reference standards.

PPVs and NPVs are also shown in Table 3. Against the MRS, tongue swab testing had PPVs of 86.7-87.5% and sputum swab of 96.4%, with NPVs of 89.4-90.0% for tongue swabs and 96.4% for sputum swabs. Against the CRS, the PPV was high (tongue swabs 93.3-95.8% and sputum swab 100%), but the NPV was low at 39.1-41.7% for tongue swabs and 50.0% for sputum swab.

## DISCUSSION

We performed the first assessment of nPOC molecular assays MiniDock MTB and MTB Ultima for childhood TB on tongue and sputum swab samples. Sputum swab MiniDock MTB had similar accuracy to sputum Xpert Ultra, meeting WHO target accuracy for an nPOC sputum molecular test. Although tongue swab accuracy was lower than sputum swab testing and the WHO TPP, it met the contextual benchmark accuracy, and MiniDock MTB had higher sensitivity than smear microscopy. Our findings demonstrate that swab-based assays have value for childhood TB to increase access to molecular testing near the point of care.

Tongue swab testing using MiniDock MTB and MTB Ultima were more accurate than previously described in children using other platforms such as GeneXpert. Xpert Ultra testing on stored tongue swabs among children from South Africa had a sensitivity of only 19% against an MRS,^11^ and 7% in prospective testing from children less than 5 years old in Kampala, Uganda.^12^ In contrast, we found that prospective tongue swab MiniDock MTB and MTB Ultima testing were 50% sensitive against a culture-based reference standard, 40% sensitive against the MRS, and highly specific. This is similar to the accuracy of MTB molecular testing of nasopharyngeal aspirates, at 46% sensitivity and 98% specificity against culture,^20^ and thus met our contextual benchmark criteria. Moreover, tongue swab MiniDock MTB had superior sensitivity to sputum smear microscopy, and the PPVs and NPVs were moderate to high for the culture and MRS reference standards. While the sensitivity of tongue swab testing is higher in adults (75-86%),^7,23^ this is substantially lower (53%) in adults who are smear negative,^10^ and likely a reflection of low bacillary load commonly seen in children and evident in our cohort where most were smear negative. This relationship has been seen in multiple studies in adults,^10,24–27^ and the sensitivity of tongue swab MiniDock MTB in adults reduces to a similar 39-48% if the participant had a very low Xpert semi-quantitative result.^10,23^ Thus, although tongue swab testing in children did not meet the WHO target accuracy for a non-sputum nPOC assay, it had comparable accuracy to nasopharyngeal aspirate, a currently WHO-endorsed sample type for children, and performs similarly to adults with paucibacillary disease.

Sputum swab MiniDock MTB testing in children had significantly higher sensitivity than tongue swab testing and smear microscopy, with similar accuracy to Xpert Ultra, and met the WHO target accuracy for an nPOC sputum molecular test for MTB.^4^ These results support the WHO extrapolation of adult and adolescent sputum nPOC recommendations to children,^28^ and suggest that there are advantages to sputum testing if feasible to collect in a child. A single device can perform both sputum molecular testing as well as tongue swab testing if sputum collection is not possible. MiniDock MTB also has advantages of being portable and battery capable, allowing use in decentralized, community-based settings where smear microscopy is the only available tool.

Overall, these findings are consistent with the recent WHO recommendation in adults and adolescents that sputum swab testing is preferred, and if not feasible, then tongue swab should be performed. Children should be considered similarly, supported by our results showing that the evaluated nPOC platforms can detect about half of those with culture-positive disease. At the same time, there are several implementation considerations. Most children with TB worldwide have negative microbiological testing, as reflected in the low sensitivity and NPV of swab testing against the CRS. Consequently, swab-based testing will need to be integrated into child TB treatment decision algorithms to ensure appropriate clinical diagnosis.

We conducted an independent validation of swab-based nPOC molecular TB platforms with a large, well-characterized cohort of children in two African countries across multiple standardized reference definitions. However, there are some limitations. Children were enrolled primarily from referral or inpatient facilities, and so further assessment is needed in community, lower-level settings with collection by routine health workers. We did not assess multiple swab testing to determine any added yield. With 38 cases of culture-confirmed TB, subgroup analyses were limited and a multi-study pooled assessment is needed. This includes subgroup evaluation of different respiratory sample types for sputum swab testing beyond expectorated or induced sputum. We did not compare tongue swab testing to stool testing, and so further work is needed to assess any added yield of combined swab and stool testing, and greater efforts are required to adapt swab platforms to analyze stool samples. The swab platforms only provided qualitative MTB results, and additional development is needed to examine drug resistance.

In conclusion, swab testing was feasible and offers a simple, near point-of-care solution for molecular TB testing for children. Overall, these results support the extension of current WHO guidelines on nPOC testing to children, with promotion of sputum collection and tongue swab testing if sputum is unavailable. Given the large TB case detection gap in children, approaches that can reduce barriers and increase access to rapid testing and earlier diagnosis are critically needed.

## Data Availability

Individual participant data that underlie the results reported in this article, after de-identification, will be available after publication upon request, scientific review of the proposal, and data sharing agreement.

## CONTRIBUTORS

Conceptualization: DJ, CK, SY, PN, CMD, AC, HJZ, NB, EW

Data Curation: AA, JL, DB, PW, RC, HP

Formal Analysis: HP

Funding acquisition: DJ, PN, CMD, AC, NB

Investigation: DJ, AA, JL, HP, DB, PW, RC

Project Administration: DJ, CK, SY, PN, CMD, AC, HJZ, NB, EW

Writing (original draft): DJ, AA, JL, HP, DB, PW

Writing (edit): all authors

HP, RC, DJ, JL and AA accessed and verified the underlying data. All authors had full access to the data and accept responsibility to submit the work for publication.

## ACKNOWLEDGEMENTS

The authors thank the staff at our clinical and laboratory facilities who provided care for the children in this study. Additionally, the authors are thankful to the children and their parents who were willing to participate in the study and help contribute meaningful data that may help future patients with the same illness.

## FUNDING

This study was supported by the National Institute of Allergy and Infectious Diseases (U01AI152087 and R01AI190419 to AC, CMD, and PN), and the SMART4TB Consortium made possible by the United States Department of State Bureau of Global Health Security and Diplomacy. The contents of the publication are the sole responsibility of SMART4TB and do not necessarily reflect the views of the U.S. Government, consortium collaborators, or members. HJZ is supported by the SA-Medical Research Council.

## DECLARATION OF INTERESTS

The authors declare no conflicts of interest. The developers and funders had no role in design, analysis, or decision to publish the manuscript. Pluslife and Molbio provided equipment and test kits for evaluation.

## AI DECLARATION STATEMENT

Artificial intelligence was not used in the analysis or writing of the manuscript.

## Notes

### Author Declarations

The Human Research Ethics committee of the Faculty of Health Sciences, University of Cape Town, the Mulago Hospital Ethical Research Committee, the Uganda National Council on Science and Technology, and the University of California, San Francisco Institutional Review Board gave ethical approval for this work.

