## Appendix for "Swab-based molecular testing near the point-of-care for childhood tuberculosis: a prospective diagnostic accuracy study"

**Supplemental Table 1. Head-to-head accuracy of sputum- and swab-based TB testing against a culture-based standard**

| Index Test | Sputum Comparator Test | Index Test Sensitivity[a] n/N, % (95% CI) | Comparator Test Sensitivity n/N, % (95% CI) | ΔSensitivity[b] % (95% CI); p-value | Index Test Specificity[a] n/N, % (95% CI) | Comparator Test Specificity n/N, % (95% CI) | ΔSpecificity[Cc] % (95% CI); p-value |
|---|---|---|---|---|---|---|---|
| **Sputum MiniDock MTB Test (N=231)** | Xpert Ultra[d] | 24/28, 85·7% (67·3–96·0) | 21/28, 75·0% (55·1–89·3) | 10·7% (-10·9 to 32·4) p = 0·45 | 199/203, 98·0% (95·0–99·5) | 196/203, 96·6% (93·0–98·6) | 1·5% (-1·6 to 4·5) p =0·45 |
| | Smear Microscopy[d] | 24/28, 85·7% (67·3–96·0) | 11/28, 39·3% (21·5–59·4) | 46·4% (24·4 to 68·5) p < 0·001 | 199/203, 98·0% (95·0–99·5) | 202/203, 99·5% (97·3–100) | −1·5% (−3·6 to 0·7) p = 0·25 |
| **Tongue Swab MiniDock MTB Test (N=231)** | Xpert Ultra[d] | 16/26, 61·5% (40·6–79·8) | 21/26, 80·8% (60·6–93·4) | −19·2% (-41·6 to 3·1) p =0·13 | 201/205, 98·0% (95·1–99·5) | 198/205, 96·6% (93·1–98·6) | 1·5% (-1·5 to 4·5) p = 0·45 |
| | Sputum MiniDock MTB Test | 16/26, 61·5% (40·6–79·8) | 22/26, 84·6% (65·1 –95·6) | -23·1% (-43·1 to -3·0) p = 0·03 | 201/205, 98·0% (95·1–99·5) | 201/205, 98·0% (95·1–99·5) | 0·0% (-2·4 to 2·4) p = 1·0 |
| | Smear Microscopy[d] | 16/26, 61·5% (40·6–79·8) | 8/26, 30·8% (14·3–51·8) | 30·8% (9·2 to 52·4) p < 0·01 | 201/205, 98·0% (95·1–99·5) | 204/205, 99·5% (97·3–100) | -1·5% (−3·6 to 0·7) p = 0·25 |
| **Tongue Swab MTB Ultima (N=163)** | Xpert Ultra/Cobas[d] | 8/17, 47·1% (23·0–72·2) | 14/17, 82·4% (56·6–96·2) | −35·3% (-69·1 to -1·4) p = 0·07 | 143/146, 97·9% (94·1–99·6) | 141/146, 96·6% (92·2–98·9) | 1·4% (-2·0 to 4·7) p = 0·63 |
| | Smear Microscopy[d] | 8/17, 47·1% (23·0–72·2) | 5/17, 29·4% (10·3–56·0) | 17·6% (-12·6 to 47·9) p = 0·38 | 143/146, 97·9% (94·1–99·6) | 145/146, 99·3% (96·2–100) | −1·4% (−3·9 to 1·2) p = 0·5 |

a. Against a culture-based reference.
b. Sensitivity difference between the index and comparator.

c. Specificity difference between the index and comparator.
d. Xpert Ultra/Cobas and smear results were obtained from the same sputum specimen used for the sputum-swab index test, or from the sputum specimen with the closest collection date to the tongue-swab index test

**Supplemental Table 2. Concordance between MiniDock MTB tongue swab and sputum swab testing**

| | Tongue swab positive | Tongue swab negative | Total |
|---|---|---|---|
| **Sputum swab positive** | 20 | 8 | 28 |
| **Sputum swab negative** | 2 | 204 | 206 |
| **Total** | 22 | 212 | 234 |

**Supplemental Table 3. Group authorship: R2D2 TB Network and ADAPT for Kids Studies**

| Name | Affiliation |
|---|---|
| Alfred Andama | Uganda Tuberculosis Implementation Research Consortium, WALIMU, Kampala, Uganda |
| Deusdit Atwine | Uganda Tuberculosis Implementation Research Consortium, WALIMU, Kampala, Uganda |
| Cynthia Biddle Baard | Department of Paediatrics and Child Health, South African Medical Research Council Unit on Child and Adolescent Health, University of Cape Town, Cape Town, South Africa |
| Herbert Bigogo | Uganda Tuberculosis Implementation Research Consortium, WALIMU, Kampala, Uganda |
| Marcia Chiluvane | Instituto Nacional de Saúde (INS), Centro de Investigação e Treino em Saúde da Polana Caniço (CISPOC), Maputo, Mozambique |
| Catherine Cook | Institute for Global Health Sciences, Center for Tuberculosis, University of California, San Francisco, San Francisco, CA, USA |
| Natalene Gallant | Department of Paediatrics and Child Health, South African Medical Research Council Unit on Child and Adolescent Health, University of Cape Town, Cape Town, South Africa; Department of Paediatrics, Dora Nginza Hospital, Gqeberha, South Africa |
| Lauren Greenberg | Elizabeth Glaser Pediatric AIDS Foundation, Washington DC, USA |
| Phath Guambe | Instituto Nacional de Saúde (INS), Centro de Investigação e Treino em Saúde da Polana Caniço (CISPOC), Maputo, Mozambique |
| Yekiwe Hlombe | Department of Paediatrics and Child Health, South African Medical Research Council Unit on Child and Adolescent Health, University of Cape Town, Cape Town, South Africa; Department of Paediatrics, Dora Nginza Hospital, Gqeberha, South Africa |
| Sonal Jain | Heidelberg University Medical Faculty Heidelberg, Heidelberg University Hospital, Department of Infectious Disease and Tropical Medicine, Heidelberg, Germany |
| Midori Kato-Maeda | Department of Medicine, Division of Pulmonary and Critical Care Medicine, University of California San Francisco, San Francisco, CA, USA |
| Marcia Mutisse | Instituto Nacional de Saúde (INS), Centro de Investigação e Treino em Saúde da Polana Caniço (CISPOC), Maputo, Mozambique |
| Winnie Nabakka | Uganda Tuberculosis Implementation Research Consortium, WALIMU, Kampala, Uganda |
| Jascent Nakafeero | Uganda Tuberculosis Implementation Research Consortium, WALIMU, Kampala, Uganda |
| Juliet Namboowa | Uganda Tuberculosis Implementation Research Consortium, WALIMU, Kampala, Uganda |
| Gertrude Nanyonga | Uganda Tuberculosis Implementation Research Consortium, WALIMU, Kampala, Uganda |
| Naythra Naryanan | Department of Paediatrics, Division of Paediatric Infectious Diseases, University of California San Francisco, San Francisco, CA, USA |
| Mark Nicol | Division of Medical Microbiology and Institute for Infectious Diseases and Molecular Medicine, University of Cape Town and National Health Laboratory Services, Cape Town, South Africa; Marshall Centre, School of Biomedical Sciences, University of Western Australia, Perth, Australia |
| Moses Nsereko | Uganda Tuberculosis Implementation Research Consortium, WALIMU, Kampala, Uganda |
| Moorine Sekadde | National Tuberculosis and Leprosy Program, Ministry of Health, Kampala, Uganda |
| Brittney Sweetser | Institute for Global Health Sciences, Center for Tuberculosis, University of California, San Francisco, San Francisco, CA, USA |
| Kerry Van As | Department of Paediatrics and Child Health, South African Medical Research Council Unit on Child and Adolescent Health, University of Cape Town, Cape Town, South Africa; Department of Paediatrics, Dora Nginza Hospital, Gqeberha, South Africa |
| Margaretha de Vos | Heidelberg University Medical Faculty Heidelberg, Heidelberg University Hospital, Department of Infectious Disease and Tropical Medicine, Heidelberg, Germany |
| Sam Wailagala | Uganda Tuberculosis Implementation Research Consortium, WALIMU, Kampala, Uganda |
| Lesley Workman | Department of Paediatrics and Child Health, South African Medical Research Council Unit on Child and Adolescent Health, University of Cape Town, Cape Town, South Africa |
